# The effects of hypertension as an existing comorbidity on mortality rate in patients with COVID-19 A systematic review and meta-analysis

**DOI:** 10.1101/2020.11.16.20149377

**Authors:** Elena Whiteman

## Abstract

**Introduction:** Coronavirus has spread throughout the world rapidly, and there is a growing need to identify host risk factors to identify those most at risk. There is a growing body of evidence suggesting a close link exists between an increased risk of infection and an increased severity of lung injury and mortality, in patients infected with COVID-19 who have existing hypertension. This is thought to be due to the possible involvement of the virus target receptor, ACE2, in the renin-angiotensin-aldosterone blood pressure management system.

**Objective:** To investigate the association between hypertension as an existing comorbidity and mortality in hospitalized patients with confirmed coronavirus disease 2019 (COVID-19).

**Methods:** A systematic literature search in several databases was performed to identify studies that comment on hypertension as an existing comorbidity, and its effect on mortality in hospitalized patients with confirmed COVID-19 infection. The results of these studies were then pooled, and a meta-analysis was peformed to assess the overall effect of hypertension as an existing comorbidity on risk of mortality in hospitalized COVID-19 positive patients.

**Results:** A total of 12243 hospitalised patients were pooled from 19 studies. All studies demonstrated a higher fatality rate in hypertensive COVID-19 patients when compared to non-hypertensive patients. Meta-analysis of the pooled studies also demonstrated that hypertension was associated with increased mortality in hospitalized patients with confirmed COVID-19 infection (risk ratio (RR) 2.57 (95% confidence interval (CI) 2.10, 3.14), p < 0.001; *I*^*2*^ =74.98%).

**Conclusion:** Hypertension is associated with 157% increased risk of mortality in hospitalized COVID-19 positive patients. These results have not been adjusted for age, and a meta-regression of covariates including age is required to make these findings more conclusive.

**Summary:** Risk of mortality is considerably higher in hospitalised COVID-19 patients who have hypertension as an existing comorbidity prior to admission.

## Introduction

In early December 2019, the first cases of a pneumonia of unknown origin were reported in Wuhan, China. Whilst initially appearing to be a localised outbreak, centred around a seafood and wet animal wholesale market in Wuhan City, within weeks it had spread to over 200 different countries worldwide, and was declared a pandemic on 12th March [1]. As of the 19th June 2020, there were 8,577,196 coronavirus cases, and 456,269 reported deaths worldwide [2]. It has been established that the pathogen responsible for this disease is the SARS-CoV-2 virus [3], a member of the coronavirus family. The disease is now largely referred to as COVID-19. Whilst the origin of the virus remains to be identified, its symptoms have been well characterised, and these include; fever, cough, fatigue, sputum production, headache, haemoptysis, dyspnoea and lymphopenia [4]. The rapid spread of the virus and its high variability in symptoms and severity prompted rapid research into host risk factors. Several risk factors for poorer outcomes have been identified, including older age, male sex, existing comorbidities and obesity [5]. Of the existing comorbidities, hypertension and diabetes are most frequently present in COVID-19 sufferers [6]. The noticeable high prevalence of hypertension in COVID-19 infection, and the identification of the angiotensin-converting-enzyme 2 receptor (ACE2) as the viral target [7] has drawn significant interest to the involvement of hypertension in COVID-19 infection. In the present study we conducted a systematic review with meta-analysis with an aim to summarise all available primary research which has examined whether hypertension is a risk factor for increased mortality in COVID-19 patients.

## Methods

### Search strategy and Study Selection

I performed a literature search of the studies published since the COVID-19 outbreak began (2019) until June 20, 2020, with no restrictions on country imposed. We ran our searches in Ovid-Embase, Ovid-Medline, and medRxiv using the following search terms: (exp hypertension/) OR (high blood pressure.mp. or hypertension) OR (hypertensive.mp.) AND the COVID-19 search strategy suggested by NICE, see Figure S1. The Web of Science database was also searched using the following terms: Topic= (COVID-19* AND hypertension*). If inclusion criteria was satisfied, the full-text of the articles was retrieved and reviewed in its entirety.

### Inclusion and Exclusion Criteria and Data Collection

The following inclusion criteria were considered when assessing the eligibility of the identified studies: empirical studies of hospitalised patients which include 10 or more participants that: 1) included patients with confirmed COVID-19 infection who also had arterial hypertension as an existing comorbidity prior to hospital admission at the time of COVID-19 diagnosis, 2) disclosed information on clinical outcomes defined as survival or in hospital mortality, and 3) compared clinical outcomes between hypertensive and non-hypertensive patients.

### Assessment of Study Quality

To assess quality of included studies in the meta-analysis, we used The Newcastle-Ottawa Scale for cohort studies (Figure S2), and the NIH quality assessment tool for case series studies (Figure S3).

### Statistical Analysis

We extracted the number of patients in each group (hypertensive vs non-hypertensive, died vs lived) and collated these in a table. We then calculated fatality rate (FR) as a percentage of patients that died out of the total number of patients within each group (hypertensive and non-hypertensive), and report these in our results section. We used a random effect model to summarise our statistical synthesis and generate a forest plot; this model considers within subject variance, and between subject variance, which is likely to exist in our pooled sample. Risk ratio (RR), 95% confidence intervals (CI) and p-values for each study are reported, as well as an overall pooled effect size estimate.

Heterogeneity was assessed using the *I*^*2*^ statistic which estimates the percentage of variation in effect sizes that is due to heterogeneity between the studies; the higher the value of the *I*^*2*^ statistic [8], the more heterogeneity is present. For the purposes of this systematic review and meta-analysis, I will accept an *I*^*2*^ statistic of no more than 95%. We then performed a subgroup analysis according to study size, to determine if heterogeneity remained the same when smaller and larger studies were separated.

Statistical significance was assumed for p ≤ 0.05. We used STATA (StataCorp. 2019. *Stata Statistical Software: Release 16*. College Station, TX: StataCorp LLC) for all our calculations. RR is considered significant if 95% CI do not cross 1.

## Results

### Study selection

Using the described search strategy, we identified 92 studies suitable for full-text review. We excluded 73 studies because they did not meet the inclusion criteria, and included the remaining 19 studies [9—27] in the meta-analysis (Figure 1). The included 19 studies scored well on quality assessment (Table S1 & S2), and met our inclusion criteria.

**Figure 1.**
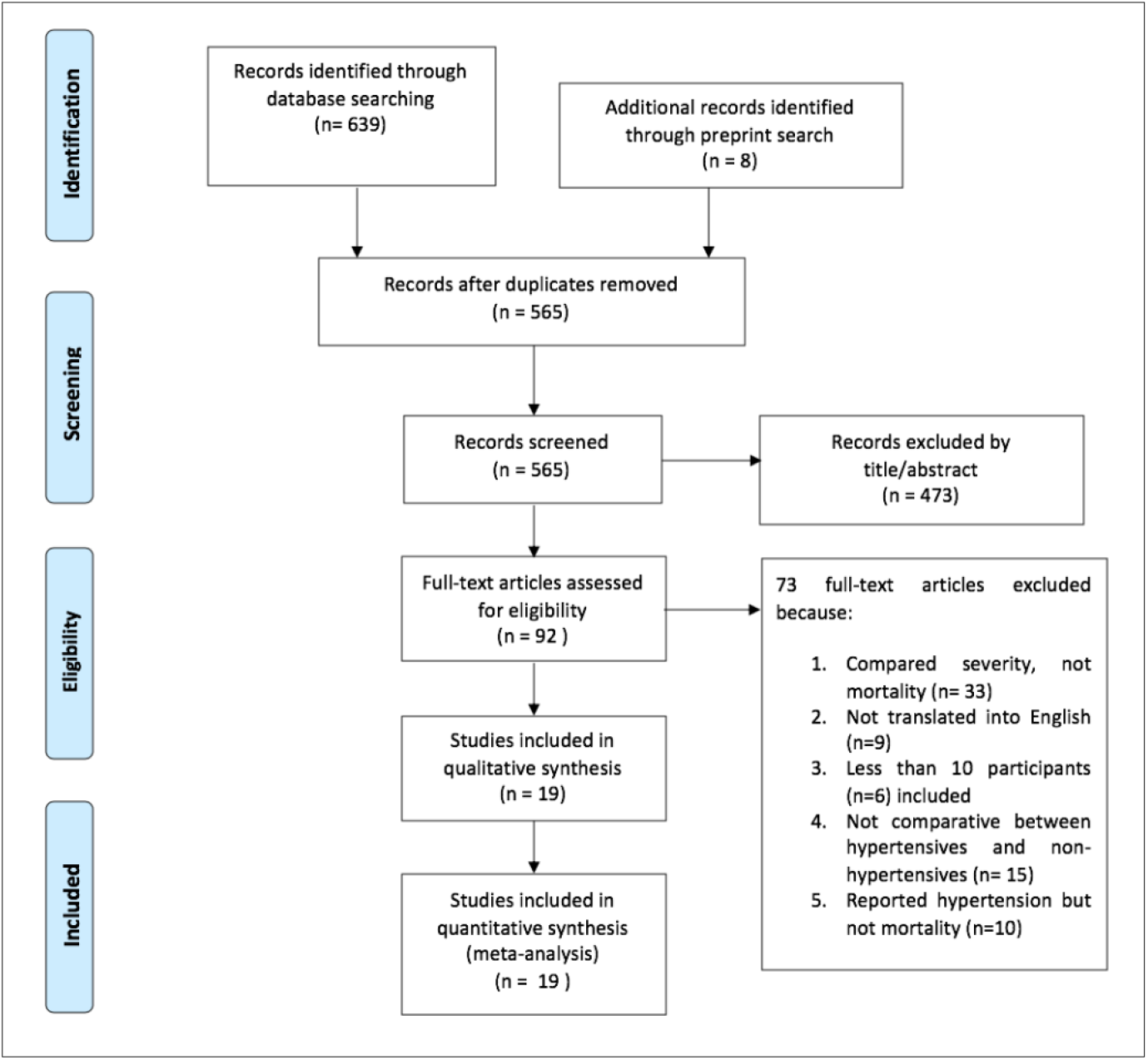
Prisma flowchart outlining study selection.

### Study characteristics

The 19 studies included resulted in a pooled number of 12,243 patients with confirmed COVID-19 infection. Of these, data on hypertension as an existing comorbidity was available for 12,243 patients, and data on survival/mortality was available for 12,218 patients. We compared mortality rates in patients with hypertension (*n*= 3,566) and patients without (*n*= 8,677). The study characteristics, including study design, hospital location, hospital admittance dates and final date of follow up can be seen in Table 1. Of the nineteen included studies, fourteen are from China, two are from Italy, two are from Iran, and one is from New York. Included studies are a mix of retrospective-cohort and retrospective case-series. All included studies define how COVID-19 was diagnosed, see Table 1, and clearly state the dates of admission for patients included in the studies; follow up period is not defined in all included studies, but ranged from 0 days to 4 weeks.

**Table 1.** Table of characteristics of included studies

| First author | Location | Study design | Sample size | Hospital admittance dates | Sampling procedure | COVID 19 diagnosis method | Final date of follow up |
| --- | --- | --- | --- | --- | --- | --- | --- |
| Akbari A et al [9] | Fars, Iran | Retrospective case-series | 440 | 18 February 2020 to 19 March 2020 | All admissions | RT-PCR assay of nasal and pharyngeal swabs | Not mentioned |
| Cao J et al [10] | Zhongnan Hospital, Wuhan, China | Retrospective case-series | 102 | 3 January to 1 February 2020 | All admissions | RT-PCR assay of pharyngeal swabs | 15 February 2020 |
| Chen T et al [11] | Tongji Hospital in Wuhan, China | Retrospective case-series | 274 | 13 January to 12 February 2020 | All admissions | RT-PCR assay of nasal and pharyngeal swabs | 28 February 2020 |
| Fu L et al [12] | Union Hospital of Huazhong University of Science and Technology, Wuhan, China | Retrospective case-series | 200 | 1 January to 30 January 2020 | All admissions | RT-PCR assay of pharyngeal swabs | Not mentioned |
| Guan WJ et al [13] | 575 hospitals across mainland China | Retrospective case-series | 1590 | 11 December 2019 to 31 January 2020 | All admissions | RT-PCR assay of nasal and pharyngeal swabs | 31 January 2020 |
| Grasselli et al [14] | Lombardy ITU network (72 hospitals) Milan, Italy | Retrospective case-series | 1035 | 20 February to 18 March 2020 | Consecutive patients admitted to ITU | RT-PCR assay of nasal and pharyngeal swabs | 25 March 2020 |
| Li K et al [15] | Quarantine unit of Tongji Hospital, Wuhan, China | Retrospective case-series | 102 | 31 January to 5 March 2020 | All admissions | RT-PCR assay of nasal and pharyngeal swabs | Not mentioned |
| Luo X et al [16] | Eastern Campus of Renmin Hospital, Wuhan University | Retrospective case-series | 403 | 30 January to 25 February 2020 | All admissions | RT-PCR assay of nasal and pharyngeal swabs | 25 February 2020 |

**Table 1.** (Continued)
|  |  |  |  |  |  |  |  |
| --- | --- | --- | --- | --- | --- | --- | --- |
| Nikpouraghdam et al [17] | Baqiyatallah Hospital, Tehran, Iran | Retrospective case-series | 2968 | 19 Feb to 15 April 2020 | All admissions | RT-PCR assay of nasal and pharyngeal swabs or clinically diagnosed based on pathognomonic CT imaging features consistent with coronavirus pneumonia | Not mentioned |
| Richardson et al [18] | New York City (12 hospitals *) | Retrospective case-series | 5700 | 1 March to 4 April 2020 | Sequential patient admissions | RT-PCR assay of nasal and pharyngeal swabs | 4 April 2020 |
| Si S et al [19] | Renmin Hospital of Wuhan University, Wuhan, Hubei Province | Retrospective case-series | 671 | 1 January to 23 February 2020 | Consecutive patient admissions | RT-PCR assay of nasal and pharyngeal swabs | 23 February 2020 |
| Tedeschi et al [20] | 10 hospitals across Italy | Retrospective cohort | 609 | 22 February to 4 April 2020 | All admissions | RT-PCR assay of nasal and pharyngeal swabs | Not mentioned |
| Wang L et al [21] | Renmin Hospital of Wuhan University, Wuhan, China | Retrospective cohort | 339 | Jan 1 to Feb 2020 | All admissions | RT-PCR assay of nasal and pharyngeal swabs | 4 weeks |
| Yan Y et al [22] | Tongji Hospital, Wuhan, China | Retrospective cohort | 193 | Jan 10 to Feb 24 2020 | All admissions | RT-PCR assay of nasal and pharyngeal swabs | Not mentioned |
| Yang G et al [23] | Hubei Provincial Hospital of Traditional Chinese Medicine, China | Retrospective cohort | 251 | January 5 to February 22 2020 | All admissions | RT-PCR assay of nasal and pharyngeal swabs | March 3 2020 |
| Yao Q et al [24] | Dabieshan Medical Center, Huanggang city, Hubei Province, China | Retrospective cohort | 108 | January 30 to February 11 2020 | Consecutive patient admissions | RT-PCR assay of nasal and pharyngeal swabs | March 3 2020 |

**Table 1.** (Continued)
|  |  |  |  |  |  |  |  |
| --- | --- | --- | --- | --- | --- | --- | --- |
| Yuan M et al [25] | Central Hospital of Wuhan, China | Retrospective case-series | 27 | 1 January to 25 January 2020 | Consecutive patient admissions | RT-PCR assay of nasal and pharyngeal swabs | Not mentioned |
| Zhou F et al [26] | Jinyintan Hospital and Wuhan Pulmonary Hospital China | Retrospective cohort | 191 | Dec 29 2019 to Jan 31 2020 | All admissions | RT-PCR assay of nasal and pharyngeal swabs | Not mentioned |
| Zhou X et al [27] | Wuhan Fourth Hospital, Wuhan, Hubei Province, China | Retrospective case series | 110 | January 25 to February 20 2020 | All admissions | RT-PCR assay of nasal and pharyngeal swabs | February 20 2020 |

\*North Shore University Hospital, Long Island Jewish Medical Center, Staten Island University Hospital, Lenox Hill Hospital, Southside Hospital, Huntington Hospital, Long Island Jewish Forest Hills, Long Island Jewish Valley Stream, Plainview Hospital, Cohen Children's Medical Center, Glen Cove Hospital, Syosset Hospital

### Fatality rates in hypertensive vs non-hypertensive patients

Comparing fatality rates in COVID-19 patients between those that are hypertensive (*n* = 3566) and non-hypertensive (*n* = 8677), there is a much higher fatality rate in the hypertensive group (30.5%) when compared to the non-hypertensive group (9.9%), see Table 2. Seven studies [10, 14, 16, 21, 24, 25, 26, 27] also report a statistically significant p-value of less than 0.01 when carrying out a between group comparison, i.e. when comparing mortality in the hypertensive vs non-hypertensive group; only two studies report an insignificant p-value of less than 0.05 [12, 15], see Table 2. It is important to note however, that only the study by Guan WJ et al was adjusted for age and smoking status; the remaining 18 studies were not adjusted for any covariates.

**Table 2.** Mortality and survival numbers in COVID-19 positive patients

| First author | N | Patients with hypertension |  |  |  | Patients without hypertension |  |  |  | p-value | HR/Univariate OR (95%CI) |
| --- | --- | --- | --- | --- | --- | --- | --- | --- | --- | --- | --- |
|  |  | n | Died | Lived | FR% | n | Died | Lived | FR% |  |  |
| Akbari A et al | 440 | 35 | 2 | 33 | 5.7 | 405 | 11 | 394 | 2.7 |  |  |
| Chen et al | 274 | 93 | 54 | 39 | 58.1 | 181 | 59 | 122 | 32.6 |  |  |
| Cao J et al | 102 | 28 | 11 | 17 | 39.3 | 74 | 6 | 68 | 8.1 | <.001 |  |
| Fu L et al | 200 | 101 | 22 | 79 | 21.8 | 99 | 12 | 87 | 12.1 | .069 |  |
| Guan WJ et al | 1590 | 269 | 28 | 241 | 10.4 | 1321 | 22 | 1299 | 1.7 |  | 1.58 (1.07-2.3) p-value= .002* |
| Grasselli et al | 1035 | 509 | 195 | 314 | 38.3 | 526 | 114 | 412 | 21.7 | <.001 |  |
| Li K et al | 102 | 31 | 7 | 24 | 22.6 | 71 | 8 | 63 | 11.3 | .138 | 2.30 (0.75-7.03) p-value= .120 |
| Lou X et al | 403 | 113 | 60 | 53 | 53.1 | 290 | 40 | 250 | 13.8 | <.001 |  |
| Nikpouraghdam et al | 2964 | 59 | 8 | 51 | 13.6 | 2905 | 231 | 2674 | 8.0 |  |  |
| Richardson et al | 2634 | 1366 | 384 | 982 | 28.1 | 1268 | 169 | 1099 | 13.3 |  |  |
| Shi S et al | 671 | 199 | 37 | 162 | 18.6 | 472 | 25 | 447 | 5.3 |  |  |
| Tedeschi et al | 609 | 311 | 131 | 180 | 42.1 | 298 | 43 | 255 | 14.4 |  |  |
| Wang L et al | 339 | 138 | 32 | 106 | 23.2 | 201 | 22 | 168 | 11.0 | .031 |  |
| Yan Y et al | 193 | 73 | 57 | 16 | 78.1 | 120 | 51 | 69 | 42.5 |  |  |
| Yang G et al | 251 | 126 | 13 | 113 | 10.3 | 125 | 8 | 117 | 6.4 |  |  |
| Yao Q et al | 108 | 16 | 7 | 9 | 43.8 | 92 | 3 | 89 | 3.3 | .001 |  |
| Yuan M et al | 27 | 5 | 5 | 0 | 100 | 22 | 5 | 17 | 22.7 | .003 |  |
| Zhou F et al | 191 | 58 | 26 | 32 | 44.8 | 133 | 28 | 105 | 21.1 | .0008 | 3.05 (1.57 – 5.92) p-value= .0010 |
| Zhou X et al | 110 | 36 | 7 | 25 | 19.4 | 74 | 2 | 62 | 2.7 | <.01 |  |
| Total | 12243 | 3566 | 1086 | 2476 | 30.5 | 8677 | 859 | 7797 | 9.9 |  |  |

\*adjusted for age and smoking status
FR= Fatality rate
HR= Hazard ratio
OR= Odds ratio

### Overall estimation of effect size and subgrouping heterogeneity differences

This meta-analysis uses a random effects model and demonstrates that the presence of hypertension as an existing comorbidity is associated with a 157% increased risk of mortality in patients diagnosed with COVID-19 (risk ratio (RR) 2.57 (95% confidence interval (CI) 2.10, 3.14), p < 0.001; *I*^*2*^ =74.98%) (Figure 2). Fourteen of the nineteen included studies report a significant p-value of less than 0.05 and 95% confidence intervals that do not cross 1.

**Figure 2.**
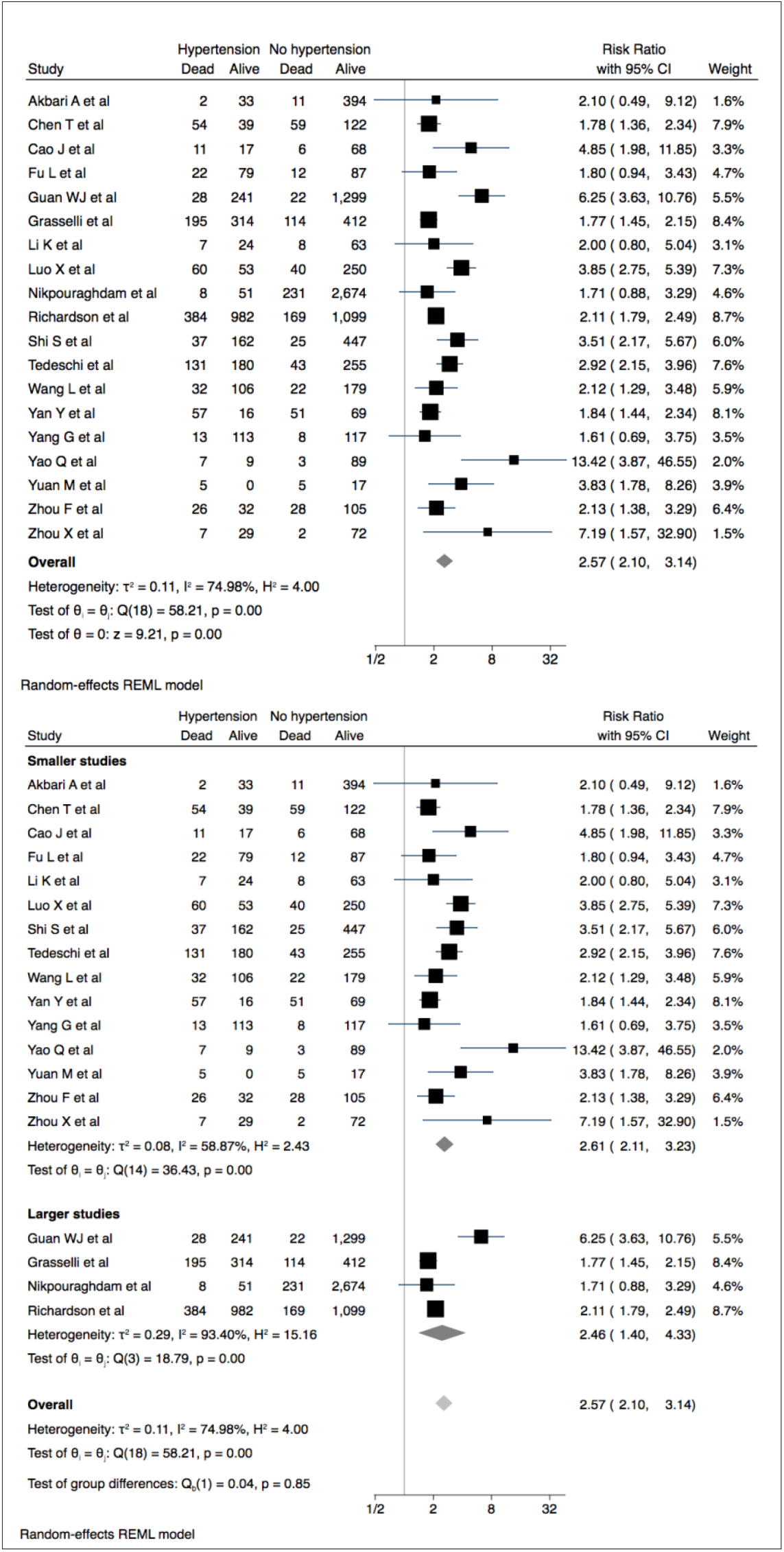
Hypertension and mortality in patients with COVID-19. Forest plot shows that hypertension was associated with increased mortality and sub-grouping larger and smaller studies demonstrates increased homogeneity in smaller studies

Overall heterogeneity in the random effects model was high (*I*^*2*^ = 74.98%), suggesting that the effects of hypertension on mortality are not the same across all studies. Subgrouping studies according to size however, Figure 2, demonstrates that heterogeneity is highest in the larger studies (*I*^*2*^ = 93.40%) and greater homogeneity is present in the smaller studies (*I*^*2*^ = 58.87%). Of the four larger studies, only one crosses the line of no difference; this is in contrast to the smaller studies, of which four of fifteen cross the line of no difference.

### Age and gender distributions within hypertensive patients

Five studies [13, 14, 20, 23, 27] provided information on age within hypertensive and non-hypertensive COVID-19 patients; three reported a median value [14, 20, 23], and two reported a mean [13, 27], see Table 3. Information on age was available for 3595 patients. Average age of patients with hypertension (*n*=1251) in the three studies reporting a median value was 69, and 62 in non-hypertensive patients (*n*=2344) (Table 3). Average age of COVID-19 positive patients with hypertension the two studies reporting a mean value was 64, and 50 in the non-hypertensive patients (Table 3). Age is therefore higher within the hypertensive patients. Although, only the study by Guan WJ et al adjusted their results for age within hypertensive and non-hypertensive patients. This study reported a hazard ratio of 1.58 (95%CI = 1.07-2.3, p-value = 0.002) when comparing mortality in hypertensive vs non-hypertensive patients, when adjusted for age and smoking status.

**Table 3.** Included studies reporting on age within hypertensive and non-hypertensive COVID-19 patients

| First author | N | Patients with hypertension |  |  | Patients without hypertension |  |  | p-value |
| --- | --- | --- | --- | --- | --- | --- | --- | --- |
|  |  | Age (years) |  |  |  |  |  |  |
|  |  | n | Measurement type | Value | n | Measurement type | Value |  |
| Guan WJ et al | 1590 | 269 | Mean (SD) | 62.1 (12.5) | 1321 | Mean (SD) | 46.2±15.6 |  |
| Grasselli et al | 1035 | 509 | Median (IQR) | 66 (60-72) | 526 | Median (IQR) | 62 (54-68) |  |
| Tedeschi et al | 609 | 311 | Median (IQR) | 76 (67–83) | 298 | Median (IQR) | 57 (47–68) | <.001 |
| Yang G et al | 251 | 126 | Median (IQR) | 66 (61-73) | 125 | Median (IQR) | 66 (57-76) | 0.685 |
| Zhou X et al | 110 | 36 | Mean (SD) | 64.8(10.1) | 74 | Mean (SD) | 54.3(14.8) | <.001 |
|  |  |  | Median average | 69 |  | Median average | 62 |  |
|  |  |  | Mean average | 64 |  | Mean average | 50 |  |
| Total | 3595 | 1251 |  |  | 2344 |  |  |  |

Information on sex was available for 2548 patients from four studies [13, 20, 23, 27]. The male sex predominated in both the hypertensive and non-hypertensive patients, at 62.5% and 57.2% respectively. Between group analysis was however, only available for two studies [23, 27] and this demonstrated insignificant results for both studies, Table 4, suggesting the sex differences in hypertensive and non-hypertensive patients may not be significant.

**Table 4.**
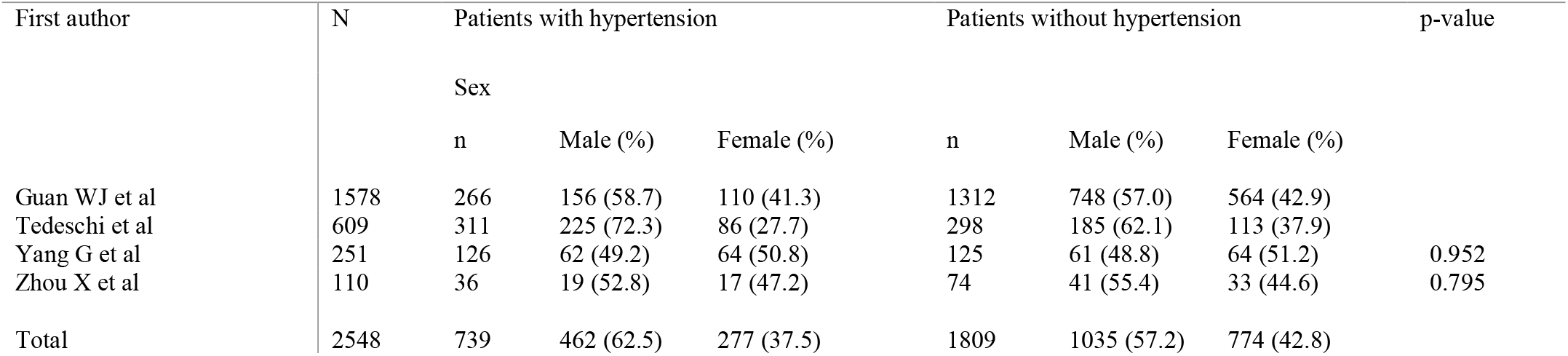
Included studies reporting on gender within hypertensive and non-hypertensive COVID-19 patients

## Discussion

### Summary of main findings

We present data that indicates a significant increase in mortality in COVID-19 patients with hypertension as an existing comorbidity; all the included studies demonstrate a higher fatality rate in the hypertensive group when compared to the non-hypertensive group, and almost all report a statistically significant between group comparison with a p-value of less than 0.05. The two studies that do not [12, 15] are both very small studies, with only 201 and 102 patients each respectively. Bias within the study, or errors in data collection could therefore easily explain the disparity in results. An overall effect estimation also demonstrates a very significant overall contribution of hypertension to increased mortality in COVID-19 patients, with only five of the total nineteen included studies crossing the line of no difference and suggesting an insignificant result. Only one of these studies [17] is within the larger studies group; the remaining four studies [9, 12, 15, 23] are all within the smaller studies group, suggesting that within study biases could be to blame for the difference in results. The smaller studies do however, demonstrate a much greater homogeneity in results when compared to larger studies, which demonstrate very high heterogeneity.

Those studies that do report on age and gender within COVID-19 patients indicate that hypertension is more commonly present in males, and in older populations. Hypertension is known to be more common in males, and the elderly, although sex differences in prevalence of hypertension are reported to diminish after the age of 60 [28]. It is already known that elderly patients infected with COVID-19 are at an increased risk for progression to a severer form of the disease, and at an increased risk of mortality when compared to younger individuals [29]. The results of this meta-analysis would suggest that the added effects of the presence of hypertension may increase this risk even further across all age groups, and this may be specifically relevant to the elderly, who are already at an increased risk. This meta-analysis however, was not adjusted for age as a co-variate and therefore the effects seen may be a mere reflection of the increased risk of mortality in hypertensive patients [30] that exists irrespective of COVID-19 infection; it would be necessary to perform a meta-regression to separate these variables and prove that the association seen is in fact true. Similarly, recent studies have identified that the male sex is more likely to die from COVID-19 infection [31]: the results of this systematic review identify that the percentage of males in both hypertensive and non-hypertensive groups, in included studies that do report on sex difference, demonstrate that males are higher in both groups. We have already mentioned that hypertension is known to be higher in males; the high prevalence of males in both the hypertensive and non-hypertensive COVID-19 patients however may suggest that these variables are indeed separate. Only four studies report on COVID-19 sex differences, and none report differences in mortality that are adjusted for sex. Again, a meta-regression for sex as a co-variate would be necessary to determine if the effects of hypertension are still present in the absence of sex as a potential confounding variable.

### The ACE2 receptor and hypertension in COVID-19

Whilst the association between COVID-19 pathogenesis and hypertension remains to be fully investigated, the proposed mechanisms by which this virus is interlinked with hypertension could be closely linked to the target receptor for this virus, ACE2, and its involvement in the renin-angiotensin-aldosterone system (RAAS). Internalisation of the COVID-19-ACE2 receptor complex would theoretically result in reduced expression of ACE2 on cell surfaces. This could then impede the cells ability to degrade angiotensin II, a necessary step to ensure correct blood pressure homeostasis.

Furthermore, hypertension is a known risk factor for increased mortality [32], and is known to cause myocardial injury [39]. Interestingly, whilst death from acute respiratory distress syndrome (ARDS) predominates in COVID-19 cases [33], there have been an increasing number of reports on myocardial injury also causing many COVID-19 deaths [34]. This could be due to increased myocardial demand that normally accompanies a viral illness, although there are several reports that a failure to appropriately metabolise angiotensin II may compromise cardiac function [35, 36], and we have already mentioned that the loss of ACE2 could interfere with this process. It remains to be established then, whether the combined effects of existing hypertension increasing risk of myocardial injury, together with the possible malfunctioning of angiotensin II metabolism in COVID-19 infection, together are causing increased risk of myocardial injury, and therefore increased mortality in hypertensive patients. Indeed, if one were to look at the results of the study by Chen T et al, it is clear to see patients with a previous history of hypertension dominated in the group of patients that developed acute cardiac injury (61% of patients) [37], although the differences in those that ultimately died does not seem as significant (77% vs 76%); the association remains to be explored.

Another possible consideration could be the management that severe COVID-19 patients which likely have a poor prognosis require. Patients who develop hypoxemic respiratory failure in ARDS will usually require mechanical ventilation [38], a process which requires general anaesthesia and loss of autonomous airway management. Hypertension is a known risk factor for complications in the application of general anaesthetic, with patients at an increased risk of greater swings in blood pressure than the normal population, followed by increased cardiovascular morbidity [39]. The increased risk that hypertension confers on mortality in COVID-19 patients could therefore be due to the requirements for mechanical ventilation that most patients suffering from a critical form of COVID-19 infection will require, irrespective of their hypertensive status.

Similarly, hyper-coagulability has been reported in COVID-19 patients, with one study noting the development of in-hospital deep vein thrombosis (DVT) in 23% of patients, despite anticoagulant prophylaxis [40]. This could possibly be explained by the long hospital-stays COVID-19 patients are faced with, although appropriate anticoagulant prophylaxis has been shown to reduce the chances of in-hospital DVTs down to 2% [41]. Hypertension is known to confer a hyper-coagulable state [42] through its well-known contributions to Virchow’s triad. Several cases of pulmonary embolisms in COVID-19 patients, in some cases bilateral, have been reported [43, 44], and these embolisms have been identified as the cause of death in some of these patients [45]. Whether or not hypertension is present in these patients however, remains unreported. It could be postulated that hypertension may be causing endothelial wall damage, thus contributing to a hyper-coagulable state, contributing to pulmonary embolisms and thus increasing mortality in COVID-19 via this mechanism. Again, this link remains to be further explored.

### Limitations and strengths

This meta-analysis does demonstrate a significantly increased risk of mortality in hypertensive COVID-19 patients admitted to hospital; however, only 12243 patients were included. To date, there have been 8,577,196 reported cases of COVID-19 worldwide and so this sample is not largely representative, and a much larger sample of patients would be needed to make these findings conclusive. The reason for such a small sample of inclusion, at least at present, is the lack of studies due to the novel nature of this disease; as time progresses, I expect there to be many more studies eligible for inclusion in a similar systematic review in future. A large level of heterogeneity between studies was also present, and this is likely to be due to confounding factors which were not controlled for in the studies. These factors can include smoking status, ethnicity, additional comorbidities, amongst others. There may also be significant discrepancies in how data was collected across all studies, as the studies included fail to define how ‘existing hypertension’ was classified. The follow-up period is also very limited, again owing to the novel nature of this disease, and therefore the mortality figures may be skewed; a longer follow up period would be preferred to allow a bigger catchment time to assess mortality in patients who were only recently admitted at the time of writing. Finally, COVID-19 is a worldwide pandemic but the included studies are not descriptive of all regions of the world. Notably, most of the included studies are from China, with only two from Italy, two from Iran, and one from New York.; it would be interesting to see the results of similar empirical studies from other affected areas in the world should they be published and translated in the near future. Finally, this meta-analysis did not adjust for covariates and so these results should be interpreted with caution; a similar systematic review and meta-analysis, followed by a meta-regression is needed to make these results more conclusive.

## Conclusion

These findings demonstrate that hypertension is a significant risk factor for increased mortality in COVID-19 patients. However, the potential reasons for this are also discussed, and hopefully demonstrate that the relationship between hypertension and COVID-19 pathogenesis is unclear; how the renin-angiotensin-aldosterone-system comes into play with this, if it does, is also unclear. More studies from across the globe, which are well-controlled and consider essential co-variates, are needed to ensure these results can be generalized to all populations; a greater understanding of COVID-19 pathogenesis is also required to determine how hypertension is conferring an increased mortality in affected patients.

## Data Availability

All data that is relevant to this research is available within the main text or supplementary material

## Patient and public involvement

The studies pooled in this meta-analysis all received ethical approval to waive the requirement for patient consent

---

**Figure S1.**
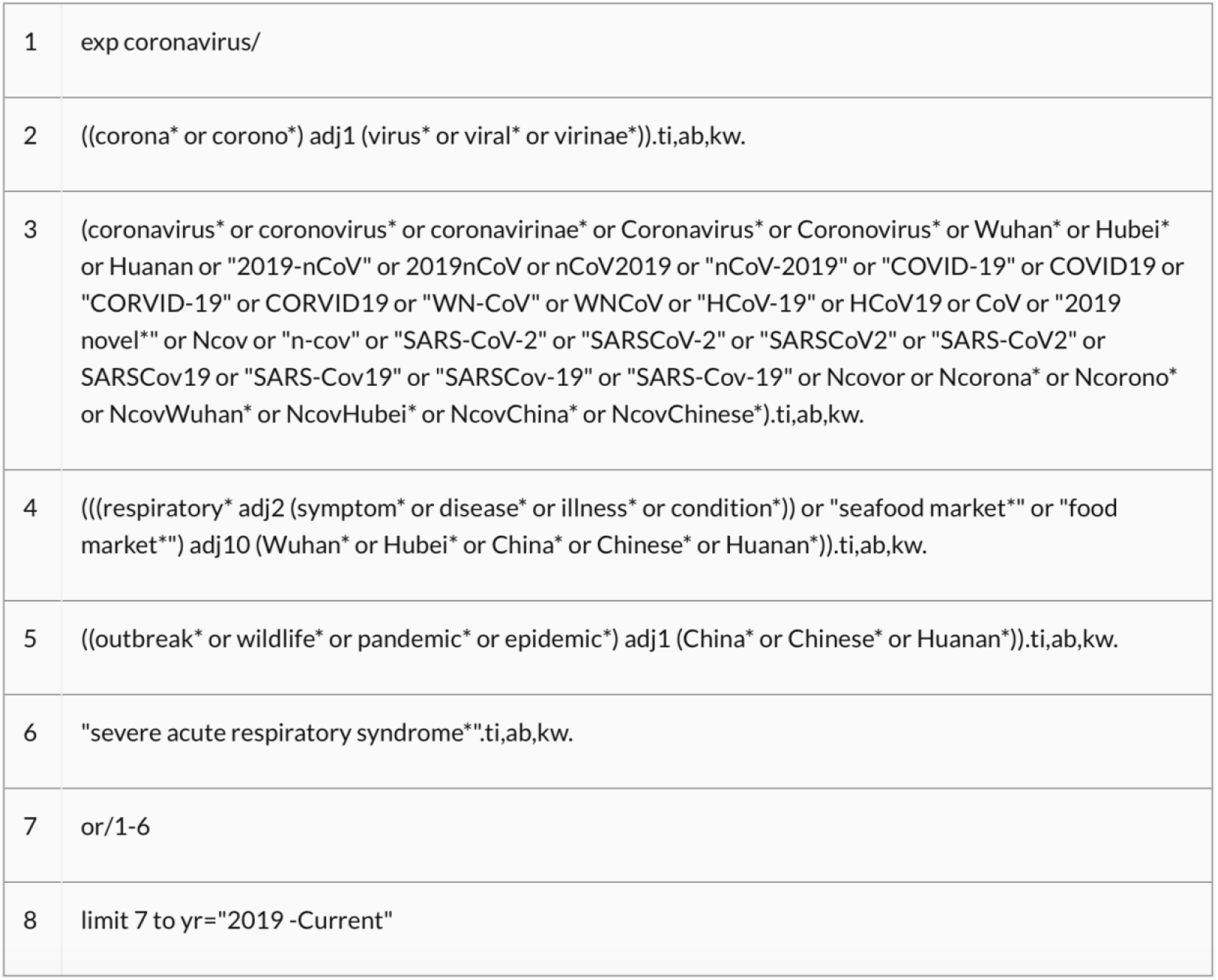
NICE recommended search strategy for COVID-19 OVID platform

**Figure S2.**
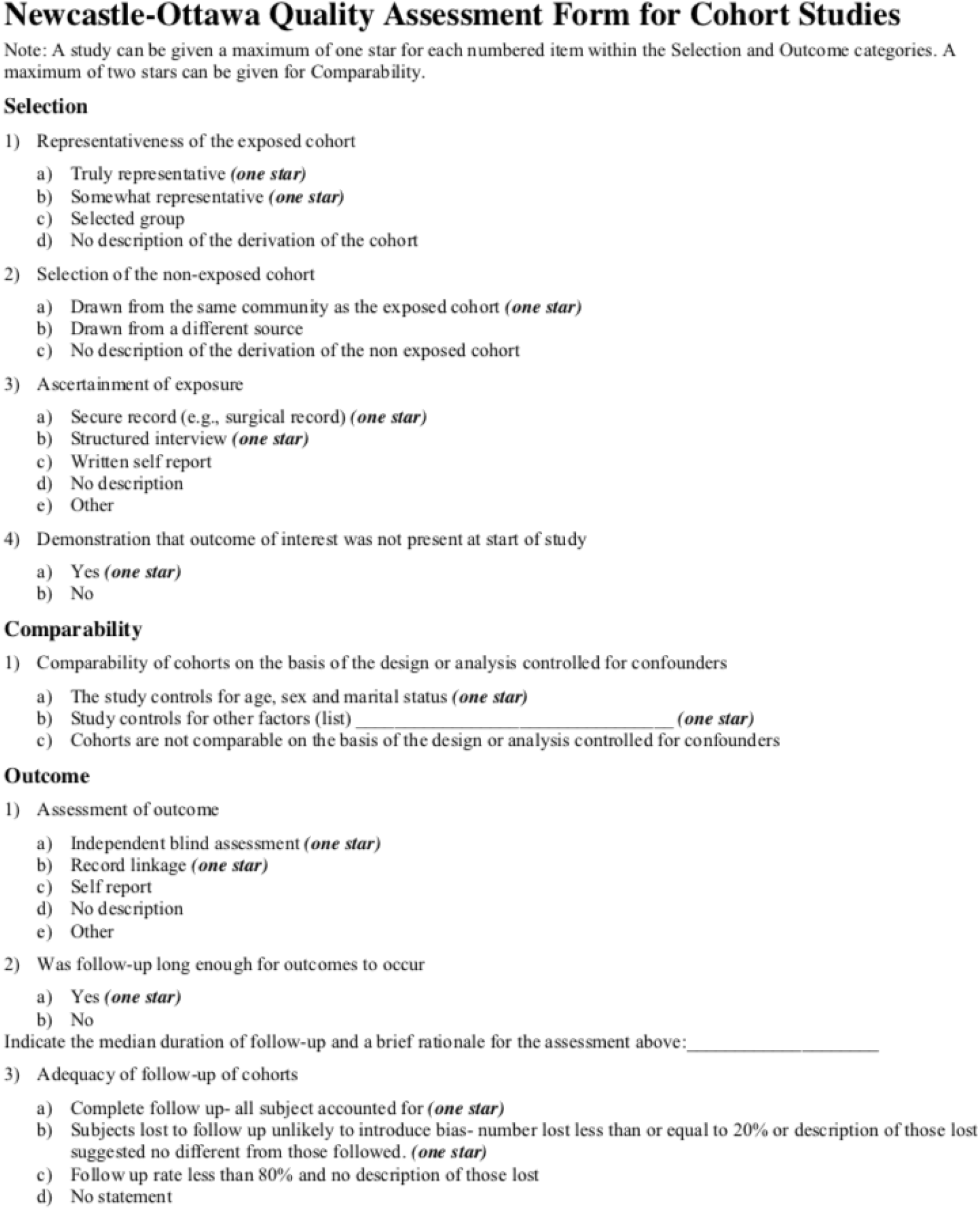
Newcastle-Ottawa quality assessment form for cohort studies used to assess quality of included cohort studies

**Figure S3.**
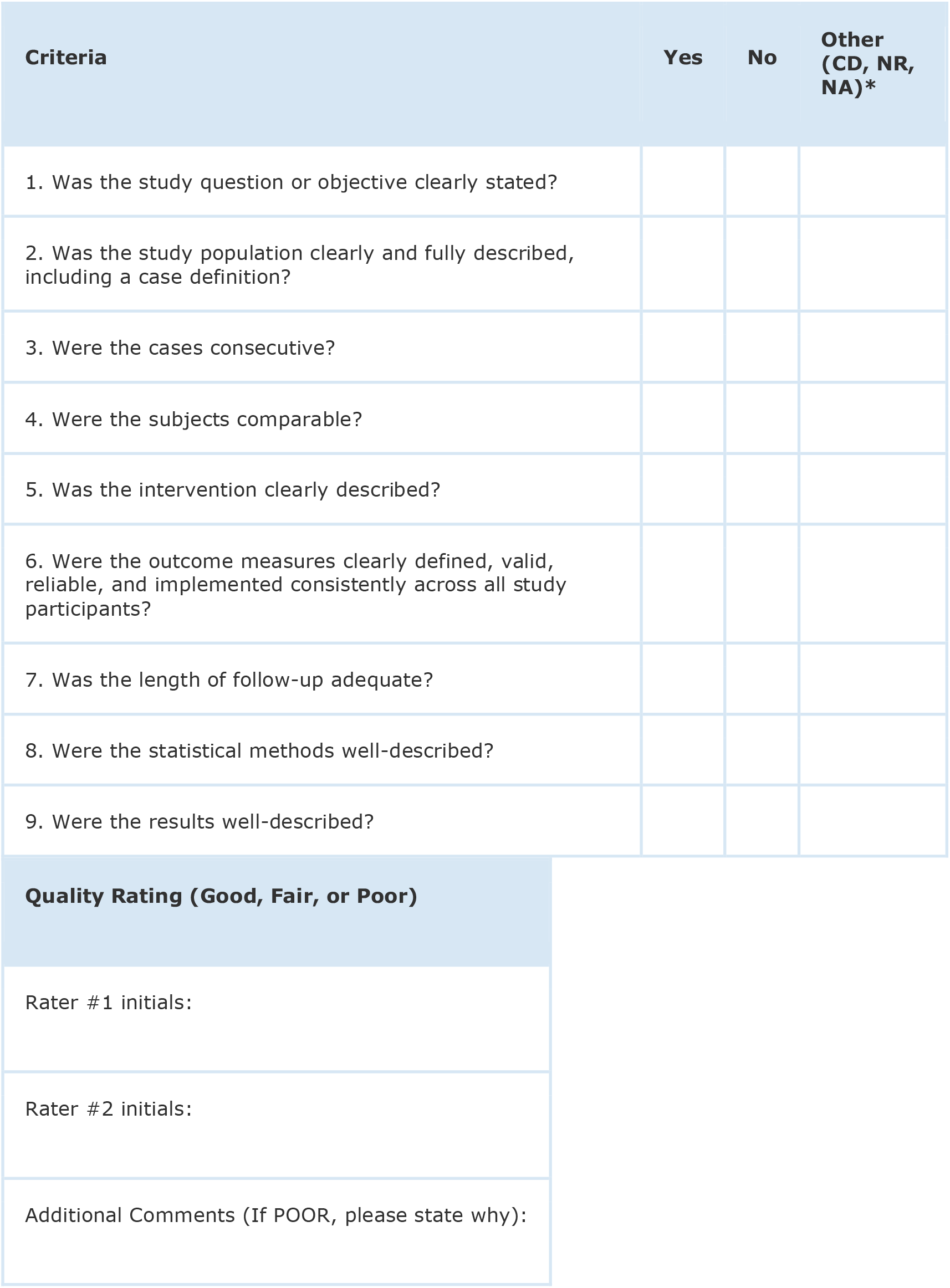
National Heart, Lung, and Blood Institute Quality Assessment Tool for Case Series Studies used to assess quality of included case-series studies *CD, cannot determine; NA, not applicable; NR, not reported

**Table S1.**
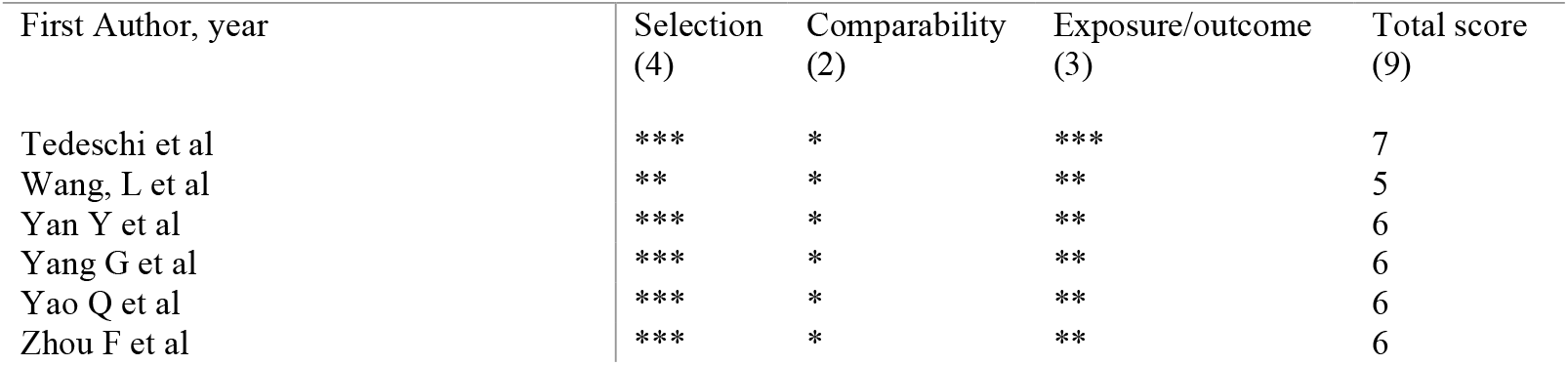
Newcastle Ottawa quality assessment for cohort studies

**Table S2.** NIH Quality Assessment Tool for Case Series Studies

| First Author, year | Quality rating (good, fair, poor) |
| --- | --- |
| Akbari A et al | Good |
| Cao J et al | Good |
| Chen T et al | Good |
| Fu L et al | Good |
| Guan WJ et al | Good |
| Li K et al | Good |
| Luo X et al | Good |
| Grasselli et al | Good |
| Nikpourghadam et al | Good |
| Richardson et al | Good |
| Shi S et al | Good |
| Yuan M et al | Good |
| Zhou X et al | Good |

